# An Analysis of SARS-CoV-2 Vaccine Breakthrough Infections and Associated Clinical Outcomes

**DOI:** 10.1101/2021.09.09.21262448

**Authors:** Heather Kerwin, Rex Briggs, Sameer Nair-Desai, Andrew Gorzalski, Mark Pandori, Stefanie Friedhoff, Thomas C. Tsai

## Abstract

Understanding the rate and clinical features associated with vaccine breakthrough infections (VBT) is of critical public health importance. Recent evidence on VBT in Barnstable County, Massachusetts, has prompted guidance on masking for vaccinated individuals in areas of high community-level transmission.^1^ Additional data is needed to better understand the prevalence and rate of VBT infections. Using detailed disease investigation data from Washoe County, Nevada we sought to assess the rate of symptomatic infection and serious illness among VBT cases compared to non-vaccinated individuals with COVID-19. From February 12 - July 29, 2021, the Washoe County Health District identified and traced 6,128 out of 6,399 reported cases across the sample period. 338 (5.5%) of all cases were identified as breakthrough infections, and 289 (86%) vaccinated individuals had symptomatic infections. Severe clinical outcomes were infrequent with 17 hospitalizations (5% of VBT) and no deaths. Cycle threshold values were not statistically different between vaccinated and unvaccinated individuals.

## [Text]

As the Delta variant causes a new surge of SARS-CoV-2 cases in the US, understanding the rate and clinical features associated with vaccine breakthrough infections (VBT) among those already vaccinated is of critical public health importance. To analyze the prevalence and features of breakthrough infections, we utilize sequencing and case data on Washoe County, Nevada from the Washoe County Health District and Nevada State Public Health Laboratory (NSPHL). Between February 12th and July 29th, 2021, the Washoe County Health District (WCHD) reported 6,399 new cases of COVID-19, and successfully traced 6,128 of these (95.8%) back to their sources (data updated as of August 3^rd^). Out of the 6,399 cases, the Health District identified 339 (5.5%) as breakthrough infections. One case was removed due to data inconsistencies, for a final N = 338. VBT cases were defined as any positive RT-PCR test for individuals fully vaccinated for 14 days or more, prior to symptom onset or routine testing. One observation was removed due to data quality, for a final sample of 338 cases. The WCHD collected individual-level data for VBT cases (N = 338), including demographics (age; sex; gender; race/ethnicity); clinical data (vaccine type and dosage schedule; symptom onset; hospitalization or death); and information on Ct-values and variants of interest, primarily from the NSPHL and, in rare instances, from other partner laboratories. Mann-Whitney U tests and Chi-squared or Fisher exact tests were used to assess differences in demographic features and outcomes associated with VBT and non-VBT cases.

Between February 12th and July 29th, VBT accounted for a small share of total infections (**Figure 1**). The 338 reported VBT cases represent 0.14% of Washoe’s vaccinated population of across the study period, compared to a 2.54% rate in those unvaccinated.

**Figure 1.**
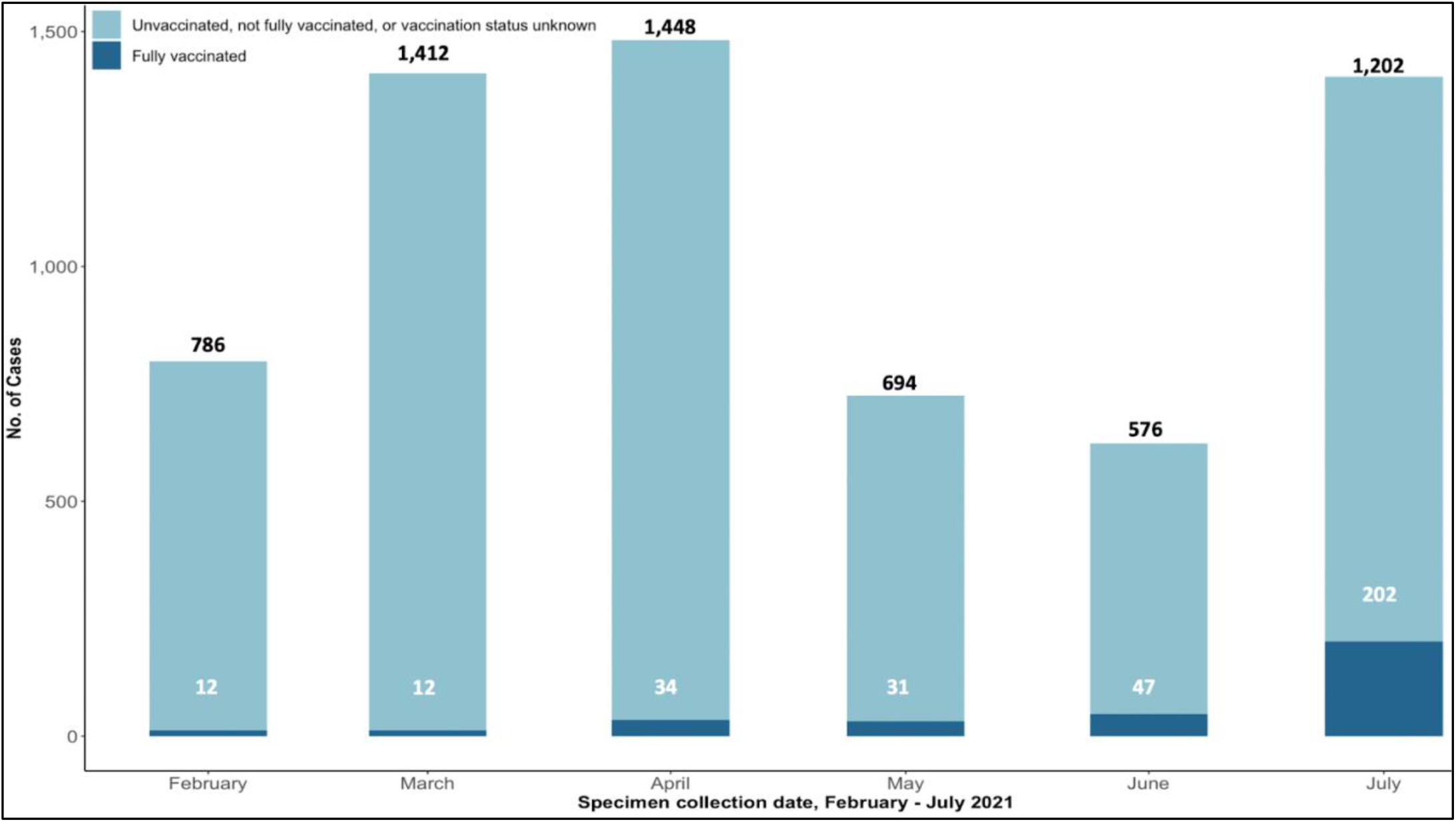
SARS-CoV-2 infections by date reported and vaccination status — Washoe County, Nevada^a^. ^a^ Calculated new cases do not perfectly align with reported cases (6,456 in Figure 1 vs. 6,399 reported) due to minor changes in county-level case reporting.

Severe outcomes in vaccinated individuals were also rare. Out of the 5,961 traced cases, 289 (86%) vaccinated individuals presented with symptomatic infections, but only 17 were hospitalized (5%) and none died. Hospitalizations mostly occurred among elderly individuals. However, elderly people were less prone to breakthrough infections and more likely to present without symptoms compared to younger age groups **(Table 1)**.

**Table 1.** SARS-CoV-2 infections and clinical outcomes by key demographics — Washoe County, Nevada.

| *Demographic Features<sup>a</sup>* | *County-Level Statistics<sup>b</sup>* | *No. of VBT Cases*<br>*(N = 338)* | *No. of Non-VBT Cases*<br>*(N = 6,061)* | *P-Values*<br>*(VBT vs. Non-VBT)* |
| --- | --- | --- | --- | --- |
| Gender |  |  |  |  |
| Male | 230,091/456,936 (50.4%) | 142/338 (42.0%) | 3,085/6,059 (50.9%) | p < 0.01 |
| Female | 226,845/456,936 (49.6%) | 196/338 (58.0%) | 2,974/6,059 (49.1%) |  |
| Age |  |  |  |  |
| 0 – 19 Years | 110,387/456,936 (24.2%) | 10/338 (3.0%) | 1,264/6,059 (20.9%) | p < 0.01 |
| 20 – 39 Years | 127,124/456,936 (27.8%) | 118/338 (34.9%) | 2,458/6,059 (40.6%) |  |
| 40 – 59 Years | 116,670/456,936 (25.5%) | 102/338 (30.2%) | 1,607/6,059 (26.5%) |  |
| 60 – 79 Years | 88,672/456,936 (19.4%) | 92/338 (27.2%) | 639/6,059 (10.6%) |  |
| 80+ Years | 14,083/456,936 (3.1%) | 16/338 (4.7%) | 91/6,059 (1.5%) |  |
| Race |  |  |  |  |
| White | 354,407/456,936 (77.6%) | 270/318 (84.9%) | 3,400/3,942 (86.3%) | p = 0.18 |
| Black | 10,630/456,936 (2.3%) | 11/318 (3.5%) | 254/3,942 (6.4%) |  |
| Asian | 24,401/456,936 (5.3%) | 20/318 (6.3%) | 245/3,942 (6.2%) |  |
| Other | 37,237/456,936 (8.2%) | 17/318 (5.4%) | 43/3,942 (1.1%) |  |
| Ethnicity |  |  |  |  |
| Hispanic | 111,653/456,936 (24.4%) | 53/317 (16.7%) | 1,564/6,059 (25.8%) | p < 0.01 |
| Not Hispanic | 345,283/456,936 (75.6%) | 264/317 (83.3%) | 4,495/6,059 (74.2%) |  |
| Outcomes |  |  |  |  |
| Symptomatic |  | 289/338 (85.5%) | 5,448/5,789 (94.8%) | p < 0.01 |
| Hospitalized |  | 17/338 (5.0%) | 411/5,789 (7.2%) | p = 0.15 |
| Deaths |  | 0/338 (0%) | 58/6,061 (1.0%) | p = 0.27 |

Out of 80 breakthrough cases sequenced from June - July 2021, 76 were associated with the Delta variant (95%). Breakthrough infections sequenced as the Delta variant had a median Ct-value of 20 (Q1 = 16; Q3 = 24; N = 77); while non-Delta breakthrough infections had a median of 21 (Q1 = 18; Q3 = 26; N = 27). Across all variants, vaccinated individuals presented similar Ct-values to unvaccinated individuals. Those fully vaccinated with an infection reported median values of 22 (Q1 = 17; Q3 = 26; N = 120), while median Ct-values for those unvaccinated were 21 (Q1 = 17; Q3 = 25; N = 797). The Kruskal-Wallis test returned a p-value of 0.8259, indicating no significant differences.

In this case series analysis from Washoe County, Nevada, the rate of breakthrough infections was more than 18-fold lower than the rate of infections among unvaccinated individuals. Most importantly, rates of severe illness were low in vaccinated individuals. These findings add to a growing body of data that can reassure the public on the real-world effectiveness of the COVID-19 vaccines.^2^

Our findings should be interpreted in light of key limitations. Early data still undergoing investigation suggests increasing rates of VBTs. Our VBT sample is not representative due to shifts in testing behavior and availability. Despite these limitations, this analysis adds to existing literature outlining the risks of VBTs and emphasizes the need for health authorities to monitor evolving trends in vaccine breakthrough data.

## Supporting information

ICJME Form for Andrew Gorzalski

ICJME Form for Heather Kerwin

ICJME Form for Rex Briggs

ICJME Form for Sameer Nair-Desai

ICJME Form for Stefanie Friedhoff

ICJME Form for Thomas Tsai

ICJME Form for Mark Pandori

## Data Availability

All data utilized in this analysis are available upon request from the lead author.

## Author Contributions

Heather Kerwin had full access to all of the data in the study and takes responsibility for the integrity of the data and the accuracy of the data analysis.

*Concept and design:* Kerwin, Briggs, Pandori, Gorzalski

*Data acquisition, analysis, or interpretation:* Kerwin, Nair-Desai, Briggs, Gorzalski

*Drafting of the manuscript:* Nair-Desai, Friedhoff, Tsai

*Critical revision of the manuscript:* Kerwin, Briggs, Pandori, Gorzalski

*Statistical analysis:* Nair-Desai, Tsai, Briggs

## Conflict of Interest Disclosures

All authors have completed ICJME forms disclosing conflicts of interest in relation to this study. No conflicts of interest were reported.

## Funding Disclosures

The Washoe County Health District and the Nevada State Health Laboratory report funding from the CDC for work unrelated to this manuscript. Thomas Tsai reports funding from William F. Milton Fund of Harvard University, the Massachusetts Consortium on Pathogen Readiness underwritten by the Massachusetts Life Sciences Center, The Commonwealth Fund, and Arnold Ventures for work unrelated to this manuscript.

## Additional Contributions

The authors would like to acknowledge Elena Varganova of the Washoe County Health District for biostatistical support and the Washoe County Health District COVID-19 disease investigators for data collection, as well as Stefanie Vanhooser from the Nevada State Public Health Laboratory for data collection and research support.

## Additional Information

All data utilized in this analysis are available upon request.

## Ethics Decision

This study has been deemed exempt from IRB/HRPP review by the Brown University Institutional Review Board, as it does not classify as Human Subjects Research.

## Notes

### Competing Interest Statement

The authors have declared no competing interest.

### Author Declarations

Brown University's Institutional Review Board did not classify our study as human subjects research, and thus IRB/HRPP review was not deemed necessary.

